# Impact of the Use of a WhatsApp-based Group Chat for Sharing Emergency Department Access-Block Data on Overcrowding and Census in a Low-Resource Emergency Department in Kigali, Rwanda

**DOI:** 10.1101/2020.06.25.20139790

**Authors:** Menelas Nkeshimana, Christine Uwineza, Amelia Y Pousson, Giles N Cattermole

## Abstract

In Low & Middle Income Countries (LMICs), hospitals often face serious communication issues that threaten to paralyze the process of healthcare provision. The Centre Hospitalier Universitaire de Kigali (CHUK) is located in the middle of a vibrant city of Kigali, and is often overwhelmed by a high number of referred patients from its catchment area, and those brought in by emergency evacuation ambulance system (SAMU). The facility has no interdepartmental landline communication network, which would be ideal to connect the inpatient services to the emergency room in order to fasten the care process. Using WhatsApp-based Group Chat for sharing the real-time caseloads, the number of patients boarding the emergency room has significantly improved (dropping from 38.1 +-7.1 to 28 +-6.5, p<0.001), although the overall length of stay in the emergency room has remained high (3.37 +-0.61 days), mainly due to other co-factors such as the availability of specialized staff (i.e. neurosurgeon) and uninterrupted imaging services (i.e. computer tomography scans).

## Introduction

### Background

Emergency Department (ED) overcrowding is a global problem that challenges clinicians in both high and low-income settings. Severe levels of emergency unit overcrowding have been linked to poor staff retention in emergency units, undesirable patient experiences in the ED, and serious increases in morbidity and mortality in high income settings and are believed to be a serious contributor to issues of low-quality emergency care in many low- and middle-income country (LMIC) settings as well (Chinonyelum 2017). As seen in other emergency care units across the world over the past several decades (Dickenson 1989), overcrowding is a shared global problem that has been associated with increased patients’ suffering, bad outcomes and a vicious cycle of emergency system malfunction (Bernstein 2009, Richardson 2006, Miro 1999, Eckstein 2004, Sun 2013, Forero 2018). In our own ED, located in a tertiary referral center in a low-income setting at the Centre Hospitalier Universitaire de Kigali (CHUK), overcrowding and access block/boarding has been linked to high rates of ED staff frustration/burnout (Pascasie 2008) and serious quality of care issues including a threefold increase in mortality for all newly admitted polytrauma cases and issues of access to care for emergency surgical patients (Petroze 2012). As in other settings, the issue of overcrowding in our emergency department has been an intractable one as many potential solutions have previously been attempted without sustained success (Byiringiro 2015, Kagobora 2014).

### Objectives

In response to this, our study represents an observational analysis of the impact of a novel pilot project using a WhatsApp-based group chat as the platform for dissemination of a daily situation report describing the level of crowding in our ED. Prior work in this setting has demonstrated severe overcrowding with high rates of >100% ED bed occupancy and very prolonged lengths of stay (Pascasie 2009, Kagobora 2014, Byiringiro 2015). The described intervention was a daily text message sent to a WhatsApp group of physician and nurse stakeholders with agency over emergency patient flow, describing the degree of overcrowding in the ED on that date.

The overarching goal of this study was to understand the impact of the use of this digital situation report (shared via a group-text on the WhatsApp application, known colloquially as the “ED sitrep”) on the real-time level of ED overcrowding. We believe this is the first report from a low-resource setting of this application of WhatsApp chat as a very low-cost solution for the ubiquitous problem of ED overcrowding. Prior estimates from high-resource settings have suggested that the time and expenses associated with the implementation of strategies to reduce emergency department overcrowding take hundreds of hours of planning and implementation and often incur sizable expenditures, ranging from USD 33,000 to 490,000 (McHugh 2012). In contrast, the ED sitrep uses a free and widely available application that is universally used in Rwanda, as in many other resource-constrained settings, for so-called “back-channel” or parallel clinical communication by clinicians (Giordano 2015, Gulacti 2016, Gould 2016, Gulacti 2017, Ganasegeran 2017, Martinez 2018). Despite the simplicity and low cost of the intervention, we have seen significant and sustained changes in overcrowding metrics in the CHUK ED and present this data as a potential model for other similar settings struggling with emergency overcrowding.

## Methods

### Study Design

The study is an ecological time-series observational trial using before-and-after comparison data related to ED overcrowding collected during roll-out of ED sitrep as a daily shared text message on a WhatsApp group chat established for this purpose. This study was approved by the hospital’s scientific & ethics committee (Ref: EC/CHUK/337/2017).

### Setting

The Kigali University Teaching Hospital/Centre Hospitalier Universitaire de Kigali (CHUK) is a tertiary referral hospital located in Kigali, the capital city of Rwanda, with a bed capacity of 500+ beds, and a catchment of 19 district and regional referral hospitals, serving directly a city of 1 million individuals and via referral, the entire population of Rwanda, totaling approximately 6 million people. The emergency department functions as a level 1 trauma & burn center, and is often profoundly overcrowded operating at >100% bed capacity the majority of the year. Historically, the 24-bed emergency department has been burdened by an entry overload with significant mismatch in terms of arrival numbers against with inpatient flow, discharges and availability of readily usable beds on the inpatient services, especially those in the high-dependency care units. The historical length of stay in the emergency room was as high as a median of 11 days, and it was very common to see patients on stretchers lining up in the hallways of the department where privacy issues limited their appropriate evaluation (Byiringiro 2015).

### Participants

As an observational trial, the unit of observation in this study was each day in the emergency department. No patient-specific data was collected or shared on the WhatsApp group as a part of the situation report. Measured variables all related to ED overcrowding and were chosen on the basis of features as described below.

### Variables, Data Sources and Study Size

The CHUK ED previously been described as facing significant challenges with overcrowding (Kagobora 2014) but did not have a central method for recording emergency department census, flow or other metrics to measure overcrowding. Patients arriving to the ED for evaluation have their demographic data, time of arrival and chief complaint recorded in a paper log-book. Thereafter their charting remains paper-based and is recorded in a chart kept at the nursing station in their walled treatment area or the nursing desk closest to their hallway bed. Prior to this study, assessments of ED volume were primarily qualitative and centered around the presence or absence of a number of patients exceeding bed capacity - during the time immediately prior to the study period the number of beds available in the four available acuity-stratified treatment areas varied from 24 to 27.

Given these challenges, for the duration of this study (Dec 16 2016-Jun 3 2017), the following procedures were followed: a daily census of the number of patients in the emergency department and the mean length of stay of those patients (LOS) was assessed and calculated by the senior consultant on duty completing a headcount and LOS assessment as a part of the daily ward-round with chart review and shared by means of a digital text-message sent to the WhatsApp SitRep group chat. On a periodic basis throughout the duration of the study, the variables from the SitRep chat were transcribed to a central electronic data repository and the ED paper log-books from the triage desk were used to calculate the volume of new patient arrivals. The log books were audited against the patients actually found to be in the ED on ward rounds and found to capture all patients pending admission and nearly all patients seen and discharged and were used as the best available estimate of ED visits per day in the absence of electronic capture of this data. The number of critical care patients arriving to the ED each day was abstracted from the paper log book used in the highest acuity area of the ED, a small ED/ICU with four monitored and ventilator-capable beds.

These variables were chosen as they were easy to document and calculate, in the interest of high levels of clinician cooperation and good data accuracy as these variables were felt to have the highest likelihood of consistency between providers while also lowering the barriers to completion given the need to hand-calculate the metrics while also supervising ongoing clinical care. Census, average length of stay and patients by service in the ED over 36 hours were used in lieu of validated scores from high-resource settings like CEDOCS and NEDOCS (Weiss 2004, Hwang 2011, Weiss 2014) as the data elements required to calculate those scores, including number of patients in the waiting room and the wait time, in hours, of the longest admitted patient were not available for data abstraction with the level of resources available in the CHUK ED. Periodic surveillance by the authors took place during this period to help ensure understanding of the definitions being used among the five clinicians contributing data to also ensure accurate data.

The group receiving the daily WhatsApp situation report included lead emergency department nurses, all ED consultants, heads or designees for all major specialist consultant services (including ICU, internal medicine, neurosurgery, orthopedic surgery, and acute care surgery), the clinical head of the emergency department and the medical director of the hospital. This daily text message from the lead ED consultant for the day included the total ED census, the mean length of stay (LOS) in days, the number of patients in the ED for more than 36 hours with the latter broken down by the admitting or consulting service responsible for the patients with prolonged LOS, hand-calculated daily after rounds as described. All texts were visible to all participants. Clinicians for the admitting services were allowed and encouraged to contribute commentary to the group regarding where they perceived blocks to exist with respect to why patients were “stuck” in the ED, ask clarifying questions and comment on their perceptions of their services’ performance when benchmarked against other services. The ED clinician was also allowed to provide answers to questions asked by consultants in addition to providing the summary data each day. In answering questions by any participant re: details of a given case, the explicit goal shared with all participants was to facilitate disposition to the wards, operating theater, counter-referral to the district level or discharge, as appropriate.

The explicit and shared objective of this situation report was to improve awareness of the severity of ED overcrowding among relevant stakeholders as this had been identified in quality improvement processes as a barrier to improvement of this as an intractable issue. The implicit and unshared objective was to use recognized natural tendencies between specialist teams to improve their own specialty’s contribution to the burden of overcrowding in comparison with others.

After the initial roll-out period from 16 Dec 2016 through 9 Jan 2017 where only census and LOS were reported, subsequent reporting periods then included a report of how many patients had been in the ED in excess of 36 hours and what inpatient clinical team was believed to be responsible for their care. A typical message reporting patients with lengths of stay over 36 hours appears below:

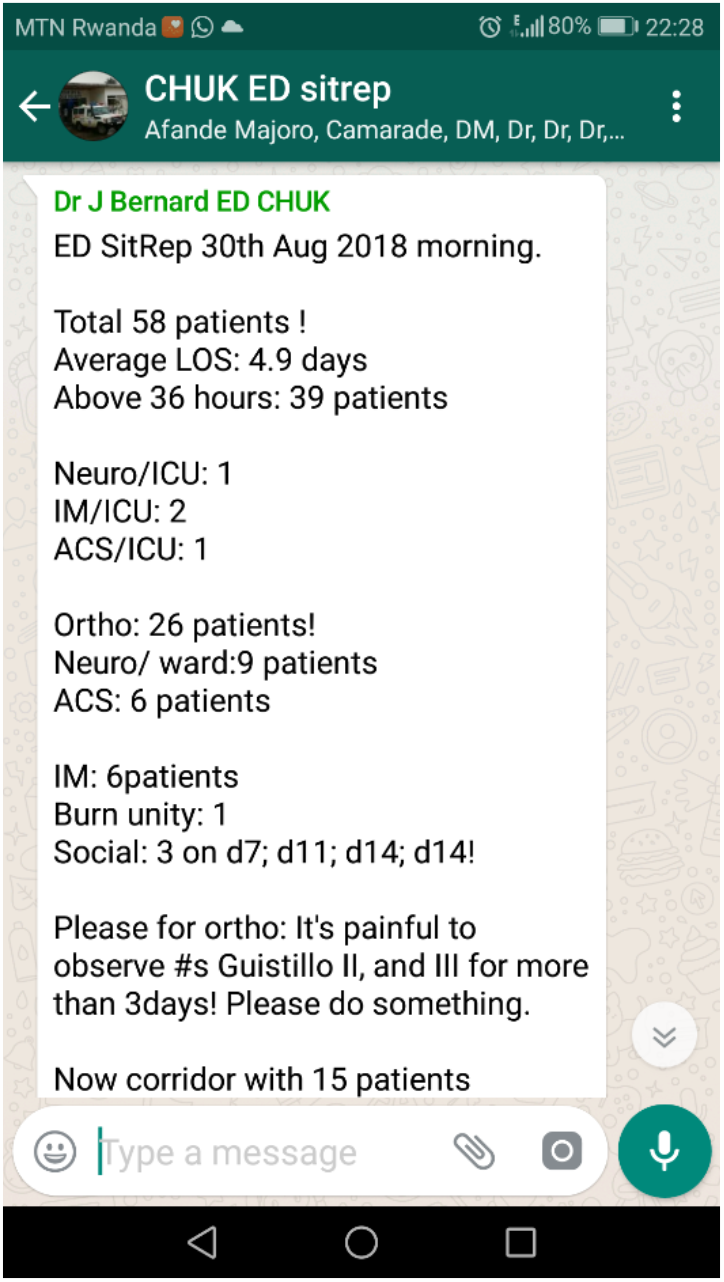

### Statistical Methods

The primary outcome of this study was to evaluate if open sharing of a minimal data set related to ED overcrowding with a group of clinician-stakeholders using the digital text-messaging service WhatsApp would decrease ED point-census as a measure of overcrowding. The secondary outcomes of this study was to evaluate if sharing of this data set would reduce the mean LOS of patients in the ED as a measure of overcrowding. Multi-way comparisons across sequential periods of observation were performed using ANOVA, pooled observations comparing the initial roll-out period where no attribution to responsible teams was shared to the period where attribution to responsible teams that followed were compared using two-sample T-testing. The number of new patient arrivals to the ED was also compared over the same sequential time periods in similar fashion to account for the possibility of a change in ED volume as a potential confounding cause of predicted reductions in census and LOS.

## RESULTS

There was a significant reduction in the total number of souls in the ED, and in their LOS, following the introduction of the WhatsApp situation report. This reduction was maintained for the duration of the study, a total of 146 days post-intervention.

To assess for the impact of confounding factors, we identified several events during the post-intervention phase that we considered might significantly affect ED census and LOS. Neurosurgical emergencies make up a large proportion of the ED caseload, and the only available neurosurgeon took annual leave a few weeks into the initial post-intervention phase. The CT scanner then stopped working, and a CT transfer arrangement was subsequently made with the private hospital. The dates and descriptions of the study phases and sub-phases are detailed in table 1.

**Table 1.** Time periods and definitions of each study phase.

| Phase | Dates | Days available | Description |
| --- | --- | --- | --- |
| 1 | 16 Dec to 8 Jan | 24 | Pre-intervention |
| 2 | 9 Jan to 31 May | 146 | Post-intervention |
| 2a | 9 Jan to 5 Feb | 28 | Post-intervention |
| 2b | 6 Feb to 1 Mar | 24 | Neurosurgeon on leave |
| 2c | 2 Mar to 31 Mar | 30 | CT scanner out of service |
| 2d | 1 Apr to 31 May | 64 | CT transfer agreement with private hospital |

Table 2 and figure 1 describe the fall in ED census and LOS between phases 1 and 2. Figure 2 shows the pattern in ED census for each of the sub-phases. Following the fall in census post-intervention (phase 2a), there was a significant rise in census during the absence of the neurosurgeon (phase 2b) which was maintained during the lack of access to CT (phase 2c). The small fall in phase 2d did not reach significance. Phase 1 census was significantly higher than each of the subsequent phases (p<0.001 throughout). Phase 2a census was significantly lower than each of the other post-intervention sub-phases (p<0.05 throughout).

**Table 2.** Souls in ED and average LOS pre- and post-introduction of WhatsApp situation report.

|  | Phase 1: pre-intervention |  |  | Phase 2: post-intervention |  |  |  |
| --- | --- | --- | --- | --- | --- | --- | --- |
|  | n (days) | Mean +/- SD | 95% CI for the mean | n (days) | Mean +/- SD | 95% CI for the mean | p |
| Souls in ED | 16 | 38.1 +/- 7.1 | 34.3 to 41.8 | 121 | 28.2 +/- 6.5 | 27.1 to 29.4 | <0.0001 |
| Average LOS | 15 | 3.84 +/- 0.60 | 3.51 to 4.16 | 120 | 3.37 +/- 0.61 | 3.26 to 3.48 | 0.006 |

**Figure 1.**
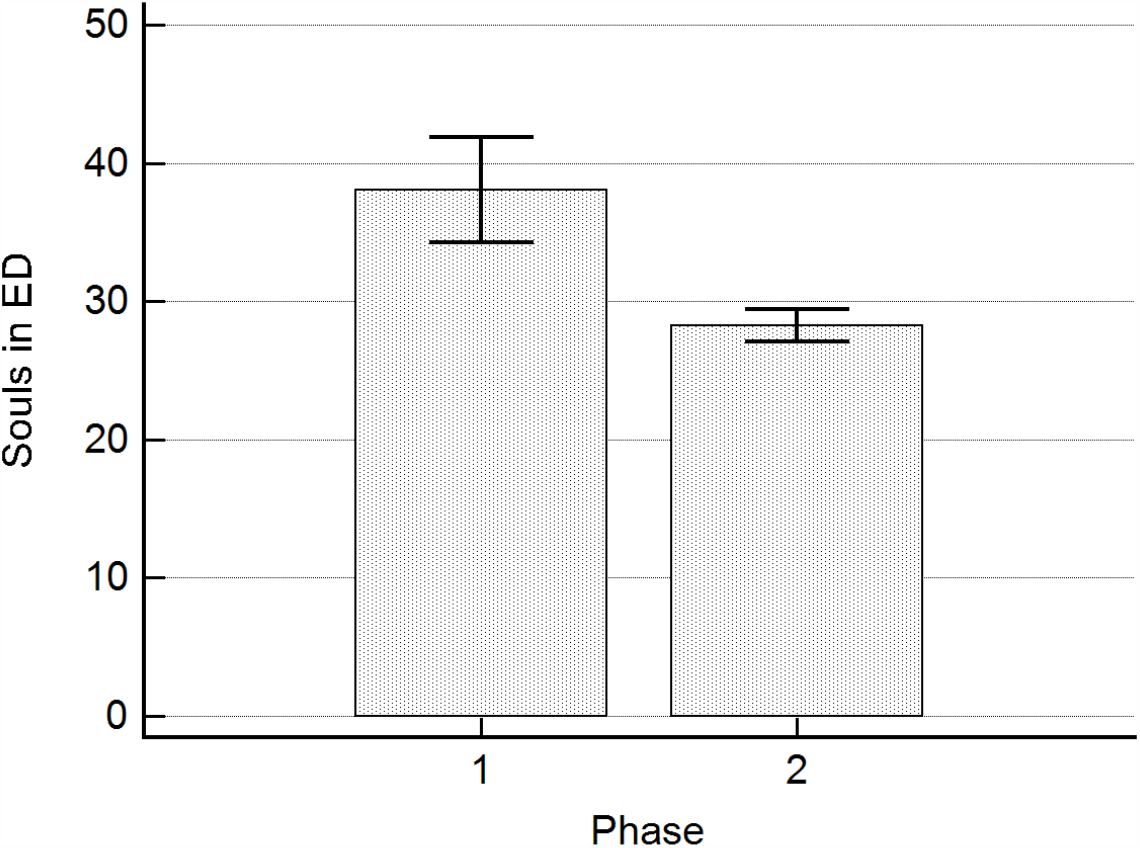
Bar chart of means +/-SD for souls in ED during phase 1 (pre-intervention) and phase 2 (post-intervention).

**Figure 2.**
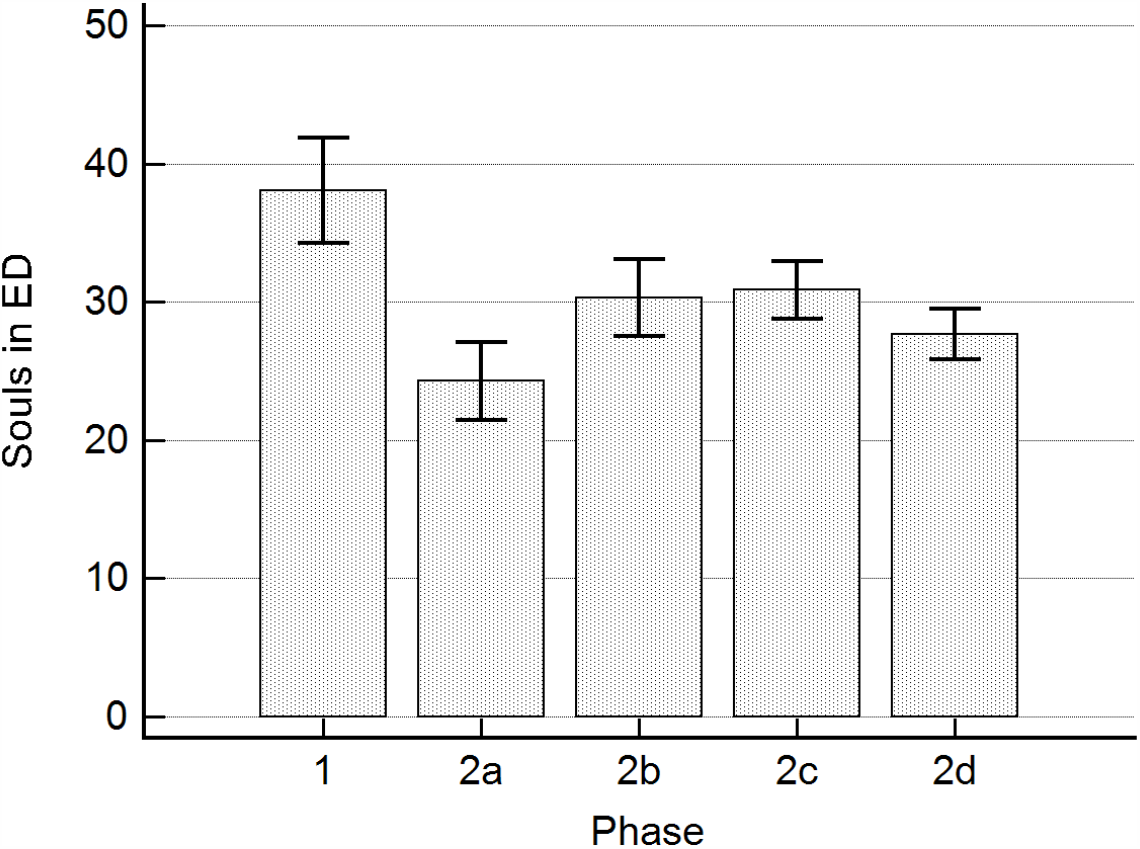
Bar chart of means +/-SD for souls in ED during phase 1 (pre-intervention) and sub-phases 2a-2d (post-intervention: see main text for explanation of sub-phases).

**Figure 3.**
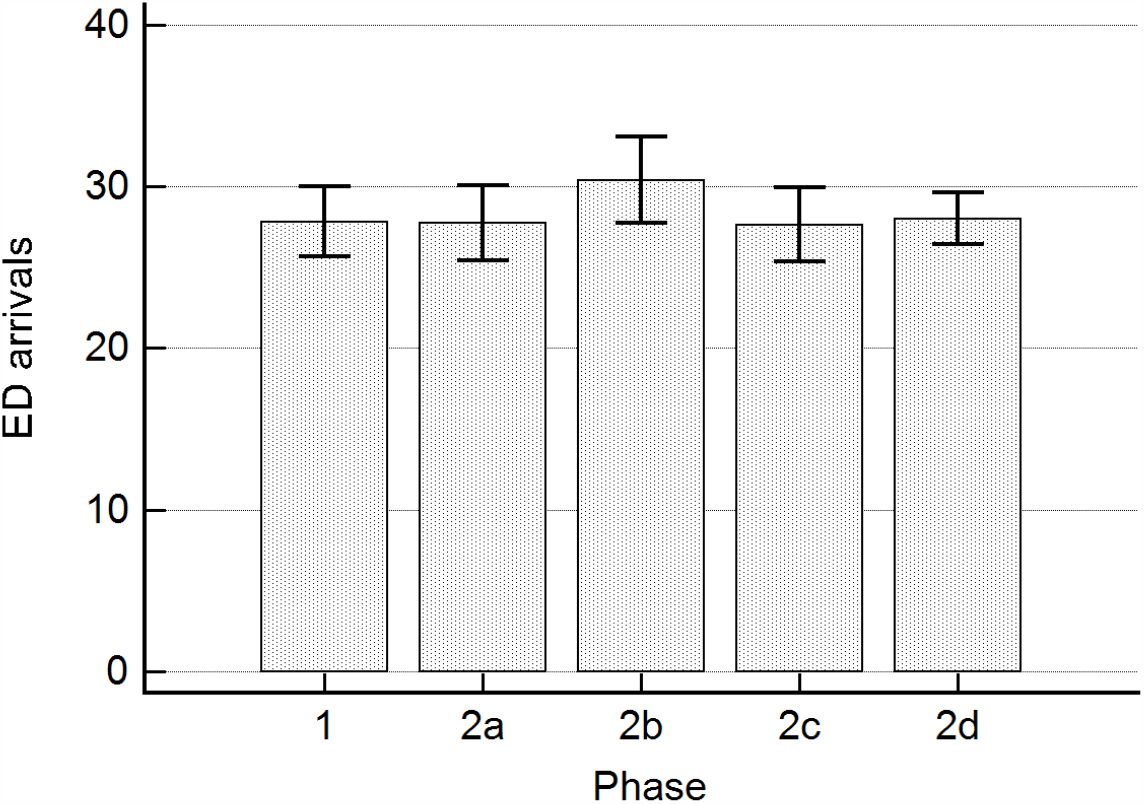
Bar chart of means +/-SD for ED arrivals during phase 1 (pre-intervention) and sub-phases 2a-2d (post-intervention: see main text for explanation of sub-phases).

- No difference in arrivals during period 1 vs 2-5 combined
- Decrease in average LOS after sitrep w >36h introduced (period 2-5)
- Decrease in average 7 AM point census after sitrep w >36h int (period 2-5)

## DISCUSSION

### Key Results

This study provides key data on the potential impact of a low-cost project using WhatsApp to improve data sharing around emergency department overcrowding in a LMIC setting. In this busy low-resource emergency department with no significant change in Emergency Department volume over the period of investigation, the introduction of a daily text-message with attribution of responsibility to services responsible for patients with prolonged LOS decreased overall census and mean length of stay for a prolonged period of several months after initiation. Despite limitations based on potential unmeasured sources of bias and design aspects related to resource constraints, these findings highlight the potential for significant gains in ED overcrowding metrics in LMIC settings at low cost and with low logistic implementation burdens.

- **No difference in arrivals during period 1 vs 2-5 combined**
- **Decrease in average LOS after sitrep w >36h introduced (period 2-5)**
- **Decrease in average 7 AM point census after sitrep w >36h int (period 2-5)**

### Interpretation

The use of the sitrep allowed a number of stakeholders with the ability to improve ED overcrowding through improvements in access-block and supported trainee-led decision making to have near real-time access to data to support decision making on patient movement and disposition. Identified strengths included the reliability of the platform in sharing the necessary data with a diversity of stakeholders to include the ED team, respective heads of consulting departments and heads of units, the user-friendliness and familiarity of the platform (WhatsApp), and the explicit shared aim of participating in data sharing and discussion with the goal of addressing the bottlenecks that were consistently leading to access block. Although the discussion did not always lead to an immediate solution to the problems identified, it has undoubtedly increased the awareness of this long-standing problem of “overcrowding & boarding the emergency room” and continued participation of stakeholders was testimony to a sense of collegial willingness to participate in the search for a long-lasting solution. In addition to the reduction in average length of stay in the emergency department as assessed in this study, comments made by participants in the WhatsApp group suggested that a change in culture resulting in a shared mental model of joint responsibility for ED overcrowding was an unplanned but welcome outcome of the SitRep chat, and it was evidenced by markedly more frequent and timely visits to the ED by the consultants (or their delegated senior residents).

The friendly interactions on the ED sitrep platform also allowed for improved efficiency in finding timely solutions to fixable problems, rather than waiting for periodic quality improvement audits and issues that would require the attention of senior management team, including issues related to infrastructure gaps, needed national expansion of number of high-dependency unit beds, hospital needs for renovation of oxygen plant, etc. were immediately flagged to be taken to the board and planned for in budgetary meetings. We believe that our hospital’s senior managers have drawn a lot of lessons from these interactions, and the problems that we are not yet able to fix today (i.e. infrastructure incompatibility etc.) will include attention to the issues underlying continued access blockage as plans evolve to remodel CHUK into a larger and more compatible hospital that will respond to our current challenges in the emergency healthcare delivery.

### Limitations & Generalizability

This study was carried out at only one of the three public tertiary academic hospitals in Rwanda, using a retrospective data pool which is tied to a number of limitations for in-depth care pathway. However, the findings from this study can be extrapolated to other centers that operate at the same level, and with similar settings in terms of logistics and infrastructures.

## Data Availability

All data related to this manuscript will be made available upon request.

## Acknowledgment

We acknowledge all the nursing staff in the department of Accident & Emergency at CHUK, and the EMCC residents’ group from the University of Rwanda for the time they dedicate to the solving of this longstanding problem of ED overcrowding.

We would like also to acknowledge support for the statistical analysis from the National Center for Research Resources and the National Center for Advancing Translational Sciences (NCATS) of the National Institutes of Health through Grant Number 1UL1TR001079.

